# Can machine learning with data from wearable devices distinguish disease severity levels and generalise across patients? A pilot study in Mania and Depression

**DOI:** 10.1101/2022.05.19.22274670

**Authors:** Bryan M. Li, Filippo Corponi, Gerard Anmella, Ariadna Mas, Miriam Sanabra, Isabella Pacchiarotti, Marc Valentí, Anna Giménez-Palomo, Marina Garriga, Isabel Agasi, Anna Bastidas, Tabatha Fernández-Plaza, Néstor Arbelo, Myriam Cavero, Clemente García-Rizo, Miquel Bioque, Norma Verdolini, Santiago Madero, Andrea Murru, Iria Grande, Silvia Amoretti, Victoria Ruiz, Giovanna Fico, Michele De Prisco, Vincenzo Oliva, Eduard Vieta, Diego Hidalgo-Mazzei

## Abstract

Mood disorders are severe and chronic mental conditions exacting high costs from society. The lack of reliable biomarkers to aid clinicians in tailoring pharmacotherapy based on distinguishable patient-specific traits means that the current prescribing paradigm is largely one of trial and error. Previous studies showed that different biological signatures, such as patterns of heart rate variability or electro-dermal reactivity, are associated with clinically meaningful outcomes. Against this backdrop, the advances in machine learning and the spread of wearable devices capable of providing continuous and ecological monitoring of patients may unlock great opportunities in mental healthcare. We herewith present a pilot study on mania and depression where we moved beyond the simple disease state binary classification but pursued the more informative and clinically meaningful task of differentiating between levels of disease severity. While most previous similar endeavours used recording segments extracted from the same subjects for both training and testing, we explicitly carried out model development and evaluation on segments from different groups of patients, in order to have a fair assessment of the model out-of-sample generalisation. This illustrated how individuals heterogeneity and non-disease-related dimensions of variations (e.g. sex, age, physical fitness) may dominate the signal so that in low sample size regimes a model might learn and overfit subject-specific patterns rather than capturing disease-relevant traits generalisable across disorders. Lastly, we developed a viable baseline for pre-processing raw data from wristband recordings and compared three classical and two deep-learning models to identify levels of disease severity.

## 1 Introduction

Mood disorders, also referred to as affective disorders, are a group of diagnoses in the Diagnostic and Statistical Manual 5^th^ edition (DSM5, American Psychiatric Association et al. 2) classification system where a disturbance in a person’s mood stands out as the dominant psychopathological feature. Disturbances in thought, energy, and vegetative functions are commonly described too. Mood disorders encompass two main diagnostic constructs, i.e. major depressive disorder (MDD) and bipolar disorder (BD). Major depressive episodes (MDE), which are characterised by feelings of sadness and loss of interest, are common in both MDD and BD, whereas BD is uniquely defined by occurrences of manic episodes (MEs). These lie on the opposite end of the mood spectrum, being characterised by increased activity and self-esteem, reduced need for sleep, expansive mood and behaviour [44]. With a lifetime prevalence of around 21% [24] and a severe, relapsing-remitting course, mood disorders constitute one of the world’s greatest public health problems, with significant direct and indirect costs, estimated at USD210.5 billion in 2010 in the United States alone [16].

Despite pharmacotherapy being generally quite effective, variability in response is poorly understood and, in the absence of biomarkers for personalising pharmacotherapy, the current medical prescription paradigm is largely one of trial-and-error [4]. In this context, and with the wide availability of wearable technology, digital biomarkers suitable to identify illness activity and provide real-time patient monitoring offer a promising approach to precision medicine in mood disorders. In fact, wearables collecting highly detailed actigraphy, sleep and cardiovascular information have already been shown to accurately capture rest-activity rhythms, illness activity and episodes in BD [14, 10, 11]. However, their longitudinal potential on treatment response and outcomes remains poorly investigated [38]. Electrodermal activity (EDA) hyporeactivity, denoting autonomic dysfunction, has long been recognized as a strong predictive biomarker for both unipolar and bipolar depression as well as suicidal behaviour [19, 15, 36]. However, until recently, capturing EDA was only possible with costly and complex laboratory equipment [13]. Novel research-grade wearables now allow to continuously record EDA in addition to 3D acceleration and heart rate variability, which are also closely linked to illness activity [40]. We herewith present a pilot machine learning implementation aimed at distinguishing the severity of different affective states of mania and depression using data collected non-interventionally with a state-of-the-art research-grade wearable in a university hospital.

In this work we investigated the potential of machine learning and wearable devices in precision medicine for mental health. Our contributions can be summarised as follows:

– Rather than settle for a yes-or-no disease identification approach, in this work we advocate for differentiation between levels of disease severity as a more clinically meaningful task in manic and depressive episodes.
– Unlike most previous works, we tested our models not only on recording samples collected from the same group of patients used for model development but also on samples from a different group of subjects to which the models have never been exposed to during training. This was to study model out-of-sample generalisation.
– We developed a viable pre-processing baseline strategy and compared the performance of different classical as well as deep learning models on the task at hand.

## 2 Related work

Previous research endeavours pursued mood states classification in BD using data from wearable devices. Côté-Allard et al. [7] introduced a deep learning-based ensemble method to distinguish manic from euthymic BD patients, which leverage long (20h) and short (5 minutes) time intervals based on actigraphy and EDA. Their classification accuracy on 47 BD patients achieved an accuracy of 91.59% in euthymia/mania recognition. Another study from the same group aimed to distinguish mania and remission in BD patients intra-individually using motor activity wearable digital data. The motor activity of 16 manic BD patients was characterised by altered complexity and variability when compared within-subject to euthymia [22]. Regarding depression versus healthy controls (HCs) identification, one study reached an accuracy of 84.00% on a sample of 23 patients with depression (both unipolar and bipolar) and 32 HCs, using motor activity data and leveraging deep neural networks and SMOTE during training for class imbalance [21]. Another study investigated differences in motor activity between 18 manic and 12 depressive inpatients with BD, and 28 HC. They reported distinctive activity patterns between inpatients with BD on a manic or depressive episode and HCs [28]. In contrast to previous works, we aim at distinguishing the severity of mania and depression, as measured at different phases. We believe that not just the mere identification but the severity quantification of mood episodes may enable better management and treatment of patients. Furthermore, while to the best of our knowledge previous studies failed to test for generalisation on samples from subjects other than the ones used for model development, we explicitly pursued this research question and showed how this can reveal a model’s shortcoming in clinical applications.

## 3 Methods

### 3.1 Study population & assessment

Four patients, two with an acute manic episode and two with an acute depressive episode, were included in this pilot. Specifically, two patients had a major depressive episode (MDE) and two patients had a manic episode (ME) in the context of BD according to the Diagnostic and Statistical Manual 5^th^ edition (DSM5, American Psychiatric Association et al. 2. The diagnosis was confirmed with a semi-structured clinical interview (SCID-5-RV. First et al. 12). Exclusion criteria included co-morbidity with another psychiatric or neurological disorder or current drug abuse. Furthermore, acute episodes of mood alterations can sometimes include symptoms of both polarities, that is to say, depressive and manic symptoms alternate or are intertwined within the same episode. Such occurrences are referred to as mixed episodes in the psychiatric nosography. We excluded any patient with mixed symptoms from this investigation as we wanted to model just the two extremes of the mood disorders spectrum, i.e. depression and mania, hypothesising that they map to distinguishable physiological patterns detectable with a wearable device. Patients were recruited at the moment of admittance to the inpatient unit of the Psychiatry ward. Over the disease course, patients were evaluated at three points in time by the same consultant psychiatrist administering the Young Mania Rating Scale (YMRS, Young et al. 47) and Hamilton Depression Rating Scale-17 (HAMD, Hamilton 17) questionnaires. YMRS and HAMD are among the most widely used scales to assess manic and depressive symptoms respectively, with a total score ranging from 0 to 60 for YMRS and from 0 to 52 from HAMD.

The three-time points corresponded to 1) `T0` acute phase (beginning of hospital admission), 2) `T1` clinically observable response onset (mid-admission), and 3) `T2` remission (soon after the patient was deemed dischargeable from hospital). Around each of the three clinical assessments, patients were provided with an E4 Empatica wristband^1^ which they were required to wear for ∼48 hours. Note that this was a non-interventional study; as such, hospital management and treatment were not informed in any way by the data collected with E4 Empatica nor was patients’ behaviour externally altered in any manner further to the requirement of wearing the wristband throughout hospitalisation. As it is the standard practice with such acute patients, they were not allowed to leave the hospital at any point of their hospitalisation. E4 devices have sensors collecting the following physiological data sampled at different sampling rates: 3D acceleration (ACC, 32Hz), intra-beat intervals (IBI, this is the time between two consecutive heart ventricular contractions), temperature (TEMP, 1Hz), blood volume pressure (BVP, 64Hz), electrodermal activity (EDA, 4Hz) and heart rate (HR, 1Hz). Note that E4 Empatica directly provides IBI as part of its output and computes it by detecting peaks (beats) of the BVP and computing the lengths of the intervals between adjacent beats. Similarly, HR is computed from IBI with a proprietary algorithm. The study was conducted in compliance with the ethical principles of medical research involving humans (WMA, Declaration of Helsinki). The assessment protocol was approved by the relevant ethical review board. All data were collected anonymously.

### 3.2 Pre-processing

The raw data from an E4 Empatica recording session comes as a collection of as many 2D arrays as there are recorded channels: ACC, BVP, EDA, HR, IBI, and TEMP. In each 2D array, columns are the device channels and rows are the recorded measurements.

Given the naturalistic setting of this investigation, despite patients being instructed to wear the wristband throughout the recording session, we noticed during exploratory data visualisation that there were instances where all measurements go to zero, i.e. the device was not detecting any physiological activity. As we were interested in physiological markers (and not potential correlations among patients’ compliance to instructions and disease state), we decided to discard instances where all channels went and stayed to zero for at least 30 consecutive seconds, as these likely resulted from patients removing the device. Removing zero-value sequences from a recording introduces gaps within it; thus, the resulting sub-recordings were handled independently during the further pre-processing steps described below.

Since sampling rate varies across different E4 Empatica channels, the raw recordings were time-aligned using the following approach. A time unit *µ* was set to one of the following values across all channels: 1, 1/2, 1/4, 1/32 and 1/64 second. If a channel’s sampling rate was higher than *µ*^−1^Hz, that channel was downsampled by taking the average value across samples within *µ*. On the other hand, if a channel’s sampling rate was lower than *µ*^−1^Hz, that channel was upsampled by padding the channel with the average value across the last sampling cycle. If a channel’s sampling rate was equal to *µ*^−1^Hz, no manipulation was applied to that channel. Furthermore, as the E4 Empatica algorithm for HR estimation needs the first 10 seconds of the signal for initialisation, HR starts being recorded with a 10-second delay relatively to other channels. Thus, in order to have all channels start at the same point in time, the first 10 seconds of channels other than HR were cropped by default.

IBI by its own definition does not lend itself to be time-aligned (this is indeed the interval between the start of consecutive ventricular contractions), furthermore, it is not usually used directly but as a stepping stone to heart rate variability (HRV) computation [39]. HRV, as the name suggests, reflects the oscillation in the time intervals between consecutive heartbeats and is a measure of the balance in the activity of the autonomic nervous system. While wrist-worn wearable devices, such as E4 Empatica, make it possible to continuously and ecologically record IBI, inconsistent IBI intervals, produced not only by ectopic beats but mainly by motion and mechanical artefacts, are well-documented phenomena with this new technology relatively to the gold standard (i.e. electrocardiography, Karlsson et al. 23). Furthermore, upon preliminary data exploration, we noticed that there were multiple instances of variable length where IBI was missing while all other channels were recording biologically plausible values. Thus, outlier (*<* 30 milliseconds (ms) or *>* 2000 ms) and ectopic (not generated within the sinoatrial node) beats were detected and marked as invalid values. Subsequently, missing values (if any) appearing at the edges of the recording (i.e. not surrounded by non-missing IBI values) were removed from the analysis and the corresponding seconds were cropped from all other channels, whereas other IBI missing values were interpolated with quadratic interpolation, which was found to be more reliable in time-series, and IBI specifically, relatively to other methods [32]. The so-pre-processed IBI was then used to derive HRV. There are several approaches to evaluate variation in heart rate, usually grouped as time domain, frequency domain and non-linear domain features. In this pilot, we extracted 1) the standard deviation of IBI (SDNN), 2) the root mean square of successive differences between normal heartbeats (RMSSD), and 3) the proportion of adjacent intervals that differ from each other by more than 50 ms (pNN50), all computed over non-overlapping consecutive 5-minute time spans; the computed value for these features was set constant on each time unit *µ* contained in the corresponding 5-minute time spans. The SDNN is predominantly (but not exclusively) indicative of sympathetic activity, while the RMSSD and pNN50 are indicative of parasympathetic activity [3]. These are popular time-domain measures and have previously been reported to be associated with mood disorders [34]. We relied on the open-source Python package `hrv-analysis`^2^ [6] to pre-process IBI. An illustration of the time-alignment pre-processing step is shown in Figure 1.

**Figure 1:**
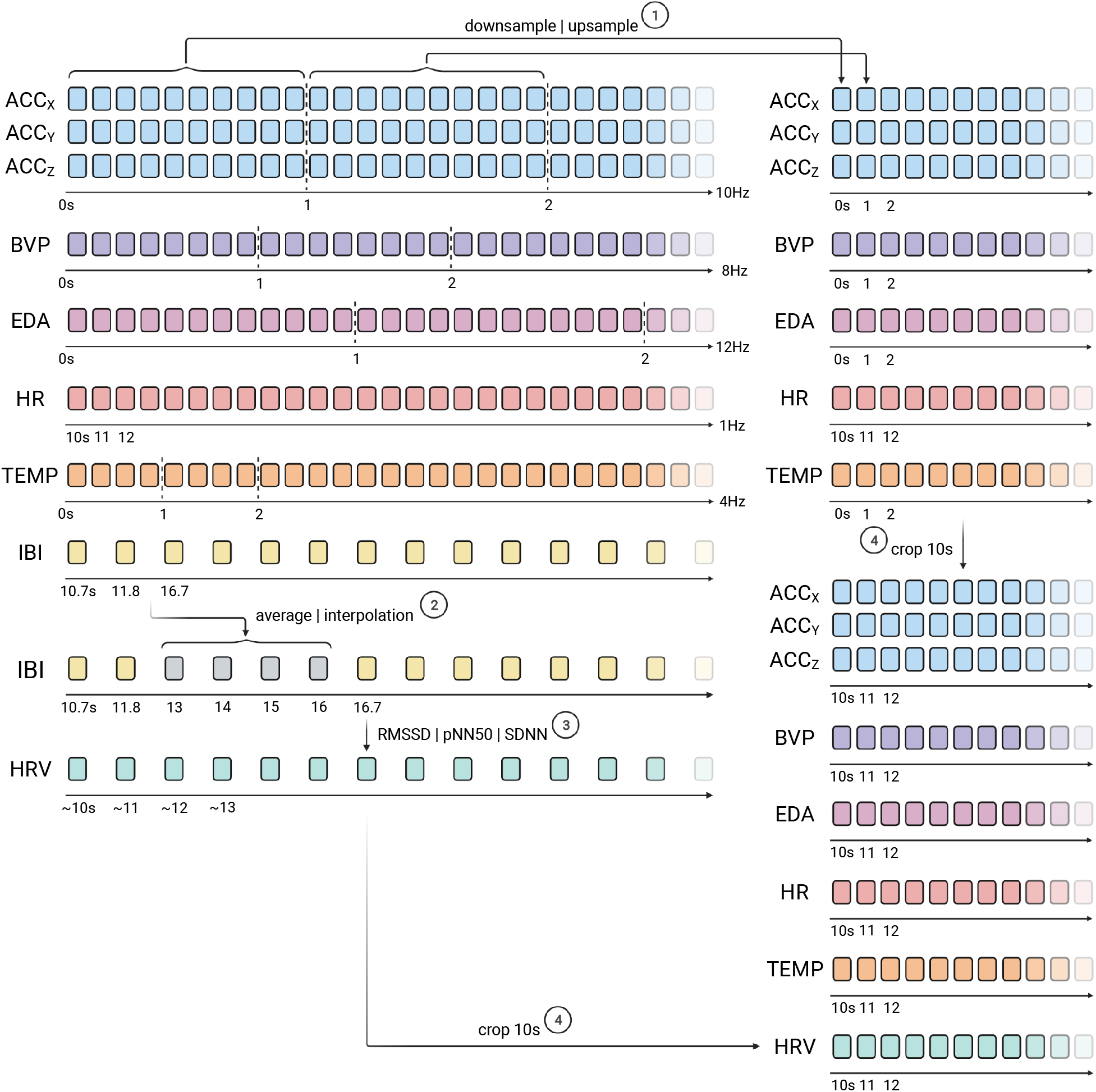
Illustration of the time-alignment pre-processing procedure. A number of variations of the pre-processing pipeline were experimented and the simplest version is illustrated here in this figure. Note that the sampling rates in the figure are made-up for illustration purposes, the actual sampling rates are specified in Section 3.1. The vertical dashed lines indicate one-second worth of recordings. The number next to each arrow indicate the order of preprocess step. Refer to the text for a fuller description of the pre-processing strategy.

Upon time-alignment following the procedure described above, each recording was then segmented into a pre-defined number of segments using a window of length *w* seconds and taking equally spaced segments with as minimal overlap between consecutive segments as possible. We explored three different powers of 2 for the window length *w*, i.e. 2^9^, 2^10^, and 2^11^ seconds (the choice of powers of 2 was motivated by computational convenience). In order to obtain an equal number of segments from each class for model evaluation, we randomly select 30 segments from each session and store them as a held-out test set to which the model is never exposed during training. We then randomly assign the remaining segments to the train and validation sets with a ratio of 80% and 20%. Each segment was finally normalised (scaled to [0, 1]) using the per-channel global (across all segments) minimum and maximum values derived from the train set. Figure A.3 shows examples of pre-processed segments (before per-channel normalisation) from the subject with MDE at different phases.

### 3.3 Experimental design

In this pilot study, we are interested in Q1) exploring to what extent a machine learning system can recognise different depressive and manic severity states using physiological data collected in a naturalistic setting with a wristband. The recording segments produced with the pre-processing steps described above were therefore used in supervised learning experiments as input to a model which was trained to classify these as belonging to one of the following states: `T0` (acute phase, at the beginning of hospital admission), `T1` (clinically observable treatment response, typically mid-admission), and `T2` (remission of the episode, last days of admission). This amounted to a six-class classification task (i.e. depression `T0`, `T1`, `T2` and manic `T0`, `T1`, `T2`). Segments from each class were extracted in the same number in order to have perfectly balanced classes. Related to the previous question, our second interest is Q2) evaluate to the model out-of-sample generalisation performance. Indeed, in a real-world deployment, we would deem a model of any value if it is able to generalise to segments drawn from unseen patients that share the same clinical condition with the subjects used for model development but that may vary widely in other respects. To this end, we divided the recruited subjects into two groups, both spanning the full spectrum of BD; that is to say, each group included one subject with MDE and one with ME. Model development was carried out with segments from one group only, while at test time we used both holdout segments from this group and segments from the other group the model had never seen during training. Generalisation estimates were computed separately for the segments from the two groups. Figure 2 illustrates the evaluation pipeline of the two groups of patients. As oppose to the train and validation sets, the proportions of segments per class and per patient were set to be perfectly balanced in the test set. Thus, accuracy was elected as the performance metric.

**Figure 2:**
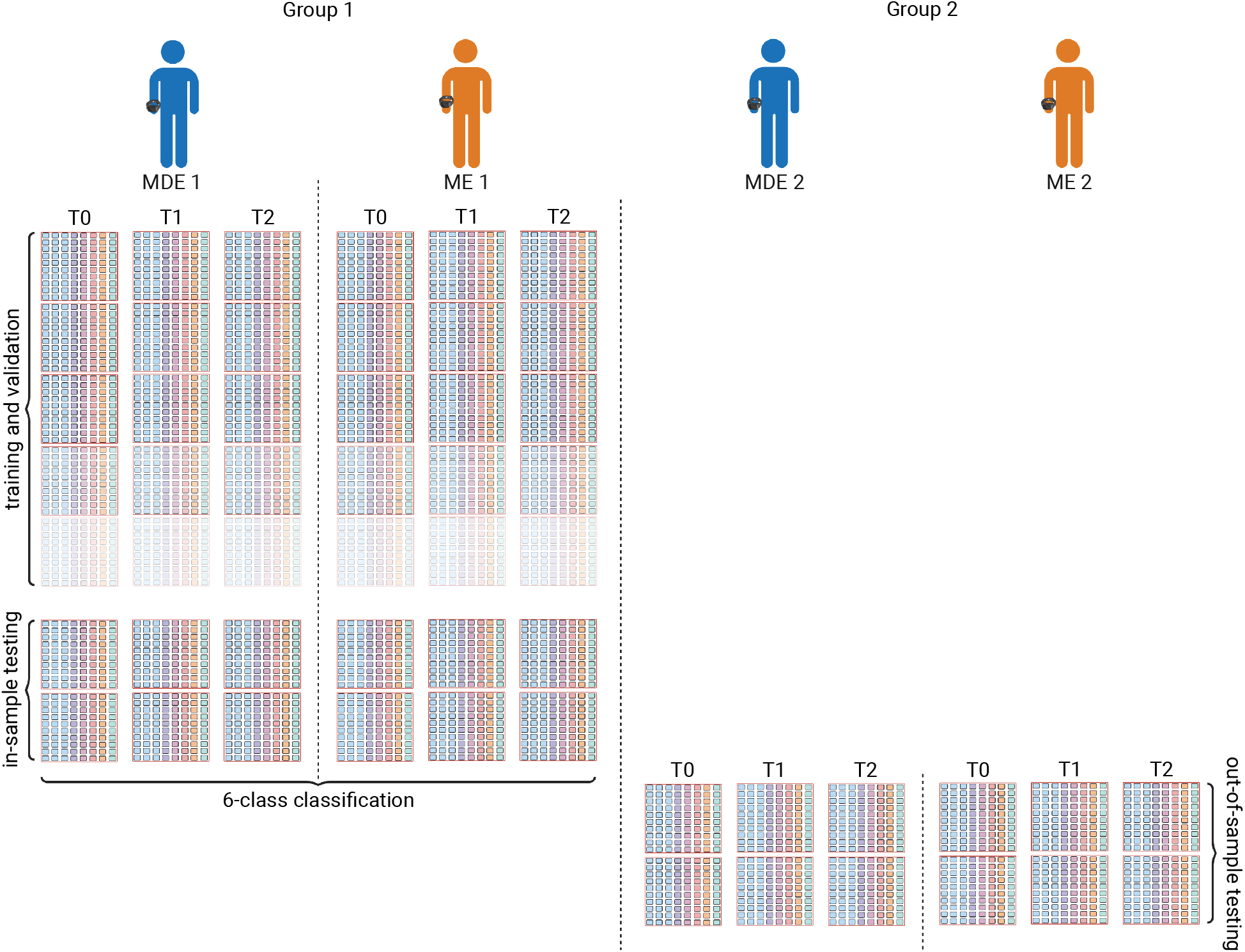
Illustration of evaluation pipeline. Two subjects with MDE (in blue, subject 1 and 3) and two with a ME (in orange, subject 2 and 4) patients were separated into two groups (subject 1 and 2 in Group 1; subject 3 and 4 in Group 2). We trained and validated the classifiers on shuffled and pre-processed segments (pink boxes, not drawn to scale) and we evaluated the trained models on the held-out test set from group 1. In addition, to evaluate the out-of-distribution performance of the models, we also test the models on pre-processed segments obtained from group 2 patients. Note that the number of segments for train and validation among different disease severity stages are not the same.

We compared three popular classic classification algorithms, i.e. 1) Elastic Net Regression (ENET), 2) Support Vector Machine (SVM) and 3) Random Forest (RF), as well as two deep neural network (DNN) models, i.e. 4) a fully-connected neural network (MLP) model and 5) a bidirectional Long Short-Term Memory (BiLSTM) model. ENET is an extension to the standard classification algorithm logistic regression adding penalties to the loss function during training with the aim of encouraging simpler models with smaller coefficient values. ENET was chosen as it is a popular method in the biomedical literature performing robustly in a wide range of applications; however, without basis functions and feature engineering, it can only fit a linear decision boundary in feature space and cannot account for interactions [30, 25]. SVM is a discriminative classifier fitting a separating hyper-plane found by maximising the separating margin between classes. It is among the most robust prediction methods, being based on statistical learning frameworks, and can efficiently perform a non-linear classification thanks to the so-called ’kernel trick’, implicitly mapping input data into a high-dimensional feature space [8]. RF is an ensemble of classification (or regression) trees, where each tree is constructed based on the principle of recursive partitioning (that is, feature space is recursively divided into regions comprising observations with similar response values). RF can learn complex, non-linear decision boundaries while still generalising well to unseen data [5]. Both SVM and RF have previously been used in similar classification tasks with data from wearable devices (e.g. Tazawa et al. 43). These non-deep-learning algorithms take as input a vector, whereas upon pre-processing our base unit of analysis was a 2D array (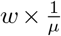 by the number of channels). So, the 2D array was flattened in order to accommodate it as an input to the aforementioned models. With the recent advancement in deep learning, DNNs have seen rapid adoption in medical research and healthcare application [31, 9]. We, therefore, also include two common DNN models in the classification experiments. The MLP model, which is fairly simplistic, consists of 3 fully-connected layers, each with Leaky ReLU (LReLU, Maas et al. 29) activation and followed by a dropout [41] layer. LSTM [18] model, on the other hand, can model recurrency in the input and is better suited for time-series and sequential data [20]. Here, we use a BiLSTM [37] model, which consists of two LSTMs, one taking the input in a forward direction, and the other in a backward direction, thereby improving the representation of temporal information in the model. DNNs can directly take 2D arrays as input data so no preliminary flattening was required here. Both DNN models were trained with Adam optimiser [26] for 400 epochs to minimise the cross-entropy between the probability distribution of belonging to any of six classes outputted by the last DNN layer and the ground-truth distribution over classes. The choice of these five popular classification models should provide a general indication of how distinguishable the severity levels are when given the pre-processed recordings.

At this stage of our project, we were interested in exploring the impact of different values of time unit *µ* and window length *w* rather than finding the optimal hyperparameters configuration for each individual model, as our focus was indeed more on the pre-processing pipeline. Thus, for each model, we evaluated a set of combinations of *µ* and *w* (15 in total), while keeping the exploration of algorithm-specific hyperparameters to a minimum, using a random search with only five draws for each model hyper-parameters configuration, the final configuration of each model is available in Table A.1.

### 3.4 Software codebase

The codebase^3^ used in this work was predominantly written in Python 3.8, where the non-deep-learning models and deep learning models were implemented using scikit-learn [33] and TensorFlow [1], respectively. In addition, all deep learning models were trained on a single NVIDIA A100 80GB.

### 3.5 Data exploration

A clinical-demographic overview of the study sample is shown in Table 1. Note that segments from the two subjects (group 2) were used at test time only, to check the model’s ability to generalise to clinically similar patients, unseen during training. While both HDRS and YMRS total scores are continuous variables, it is the usual clinical practice to bin the total score into semi-quantitative estimates of disease severity. Accordingly, the transition from `T0` (acute phase) to `T2` (remission) can therefore also be interpreted in terms of changes from severe to moderate and finally mild illness activity. Table 2 shows the number of segments per train, validation and test set used for model development (subjects in group 1). Note that this varies based on the choice of window length *w* but not time-unit *µ*.

**Table 1:** Overview of the study sample. The recording segments extracted from the subjects in the last 2 rows (group 2) were used for testing only. The recording duration shown is rounded to the closest hour. HDRS: Hamilton Depression Rating Scale total score; MDE: Major Depressive Episode; ME: Manic Episode; YMRS: Young Mania Rating Scale total score; `T0` (acute phase, at beginning of hospital admission), `T1` (clinically observable treatment response, mid-admission) and `T2` (remission of the episode, last days of admission).

| Age range (years) | Sex | Diagnosis | Hours registered |  |  | HDRS |  |  | YMRS |  |  |
| --- | --- | --- | --- | --- | --- | --- | --- | --- | --- | --- | --- |
|  |  |  | T0 | T1 | T2 | T0 | T1 | T2 | T0 | T1 | T2 |
| 56 - 60 | M | MDE | 41 | 41 | 42 | 33 | 13 | 7 | 7 | 2 | 0 |
| 26 - 30 | F | ME | 44 | 42 | 42 | 7 | 4 | 4 | 30 | 20 | 5 |
| 41 - 45 | M | MDE | 41 | 45 | 45 | 27 | 11 | 7 | 4 | 1 | 1 |
| 21 - 25 | M | ME | 48 | 45 | 44 | 3 | 5 | 4 | 23 | 15 | 1 |

**Table 2:** The number of train, validation and test segments extracted from recordings obtained from group 1 with different window length *w*. Note that we randomly select 30 segments from each recording session for the test set in the six-class classification task such that each class have an equal number of inputs in testing.

| window length $w$ | train | validation | test |
| --- | --- | --- | --- |
| **512** | 1280 | 316 | 180 |
| **1024** | 567 | 139 | 180 |
| **2048** | 213 | 49 | 180 |

In order to get a sense of the behaviour of our data upon pre-processing, we plotted and herewith report the channel pairwise normalised mutual information (NMI, Ross 35), and distribution across the entirety of the recordings used for model development. Figure 3a shows the feature distributions after per-channel normalisation. Overall, ACC, TEMP and RMSSD exhibit a relatively high level of variance, whereas the distributions of EDA, HR and SDNN sharply peaked around a single value across most recordings. Interestingly, the variance of EDA increases, and conversely, decreases in pNN50, as the condition of the subject improves (see Figure A.1a and Figure A.1c). High NMI values were found among IBI-derived HRV features, which are indeed all derived through simple transformations from IBI. High values were also recorded for ACC, TEMP and HR, which are in fact physiologically related. BVP had near-zero NMI with other channels. On a visual inspection (see Figure 3b), no major differences could be appreciated in either pairwise NMI or distribution across recordings from different classes. The feature distributions and pairwise NMI for the manic patient in group 1 are available in Figure A.2, which follow similar trends. In addition, we are also interested in knowing how informative each channel is in identifying their respective disease states. We observed that, out of the 10 features, the IBI derived features are most informative with respect to the predictive task, whereas BVP is the least informative, despite the fact that E4 Empatica derives IBI from BVP. Such result also coincides with the channel pairwise NMI. The pairwise channel-class NMI is available in Table 3.

**Table 3:**
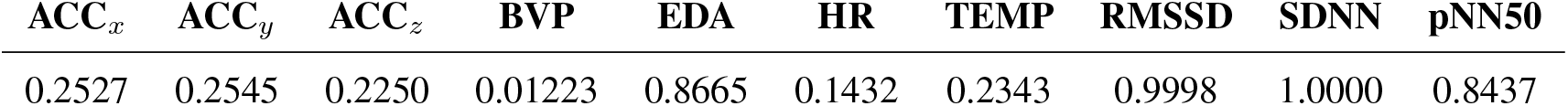
The normalised mutual information between time-aligned channels and disease states (depressive `T0` to `T2` and manic `T0` to `T2`). Note that the values below are normalised based on the maximum mutual information obtained across all channels.

| ACC<sub>x</sub> | ACC<sub>y</sub> | ACC<sub>z</sub> | BVP | EDA | HR | TEMP | RMSSD | SDNN | pNN50 |
| --- | --- | --- | --- | --- | --- | --- | --- | --- | --- |
| 0.2527 | 0.2545 | 0.2250 | 0.01223 | 0.8665 | 0.1432 | 0.2343 | 0.9998 | 1.0000 | 0.8437 |

**Figure 3:**
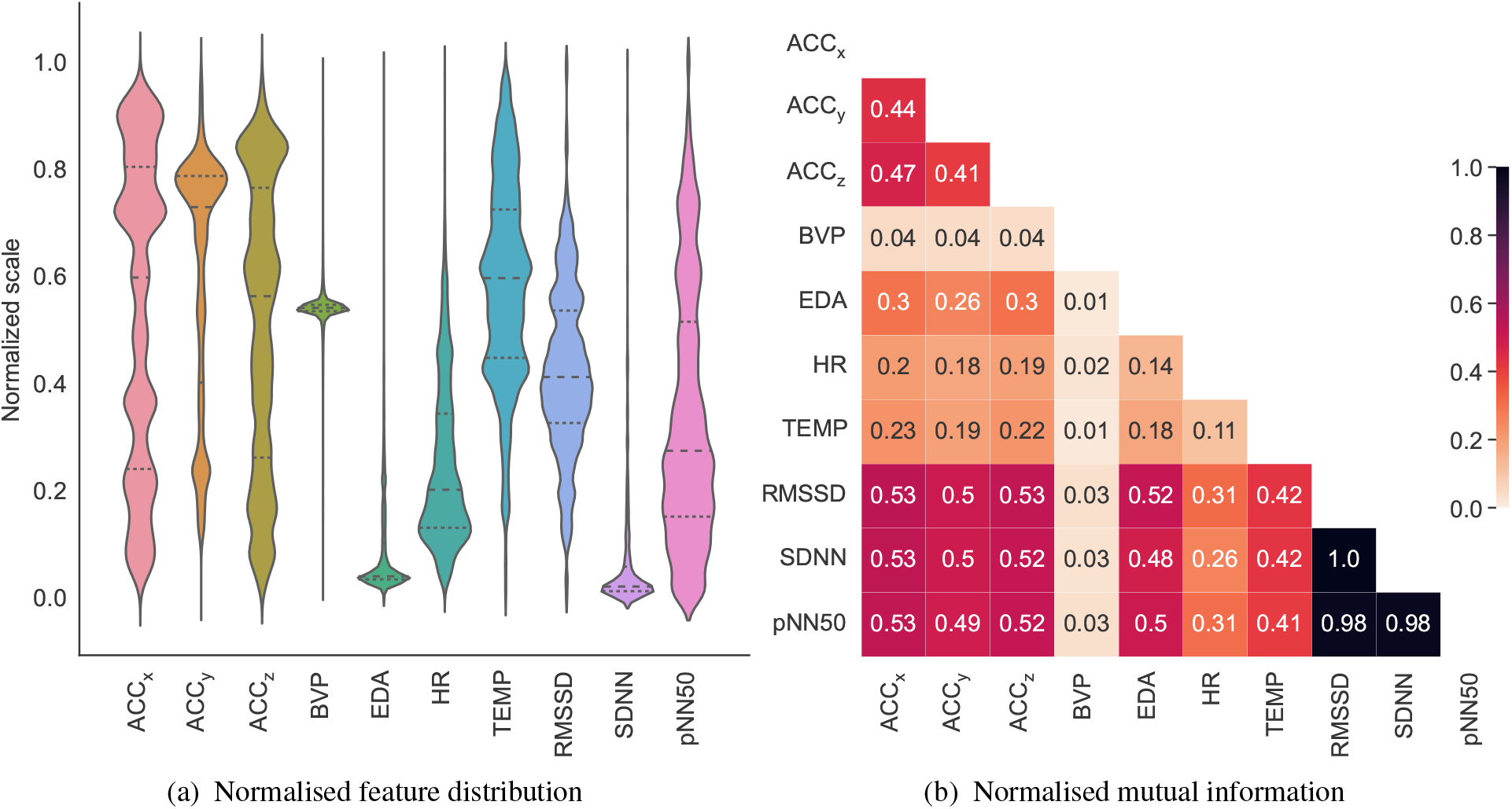
The (a) channels distributions after per-channel normalisation and (b) channel pairwise normalised mutual information of time-aligned recordings obtained from the subject on a major depression episode from group 1 at `T0`. Note that RMSSD, SDNN and pNN50 features are derived from IBI. Plots for `T1` and `T2` in the same subject as well as plots for `T0`-`T2` in the subject on a manic episode from group 1 are available in Figure A.1.

## 4 Results

During model development with segments from subjects in the first two rows of Table 1, both MLP and BiLSTM reached high accuracy in the six-class classification task on hold-out segments from the same two subjects, 68.33% and 72.78% respectively. All classical models lagged behind, with RF scoring best in this category of models (50.25%) and SVC performing worst (44.44%). However, when the trained models were deployed on segments from the two new subjects (group 2), accuracy sharply dropped across the board, with BiLSTM, the best model, at 17.78% being only slightly better than randomly guessing. Interestingly, the margin in test performance between DNNs and other algorithms significantly reduced here with ENET (the best and simplest model in its category) even outperforming MLP (see Table 4). A more fine-grained visualisation of the group 1 test performance of the two DNN models is provided by the confusion matrices in Figure A.4. For both DNN models, they were able to achieve better classification results by distinguishing the 3 depressive states (MLP: 76.13%; BiLSTM: 82.02%) as compared to the manic states (MLP: 62.92%; BiLSTM: 65.17%).

**Table 4:**
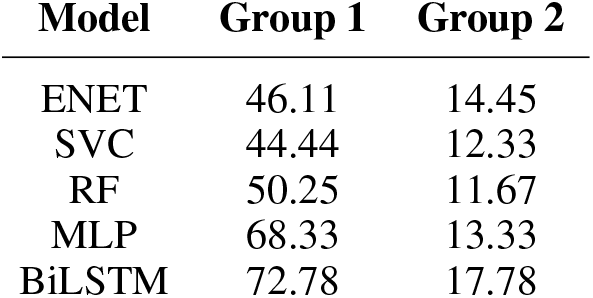
The severity levels classification accuracy of the 5 models on the (group 1) two seen patients and (group 2) two unseen patients on the held-out test set (*µ* = 1/2 and *w* = 512).

The exploration of the impact of varying *w* and *µ* on validation set accuracy showed a trend, consistent across models, whereby shorter window length *w* and, conversely, longer time unit *µ* for time alignment would, in general, be associated with an increase in validation accuracy, shown in Figure 4. By increasing the time-unit, where interpolation is needed for some channels, artificial information was introduced into the segments, hence potentially harming the models’ ability to extract relevant features. In addition, the models should be able to identify similar features in segments with longer window lengths as do in short window lengths (e.g. by ignoring the addition time-steps), though this also limits the number of training segments (see Table 2 for the dataset size) which data-hungry models such as DNNs require.

**Figure 4:**
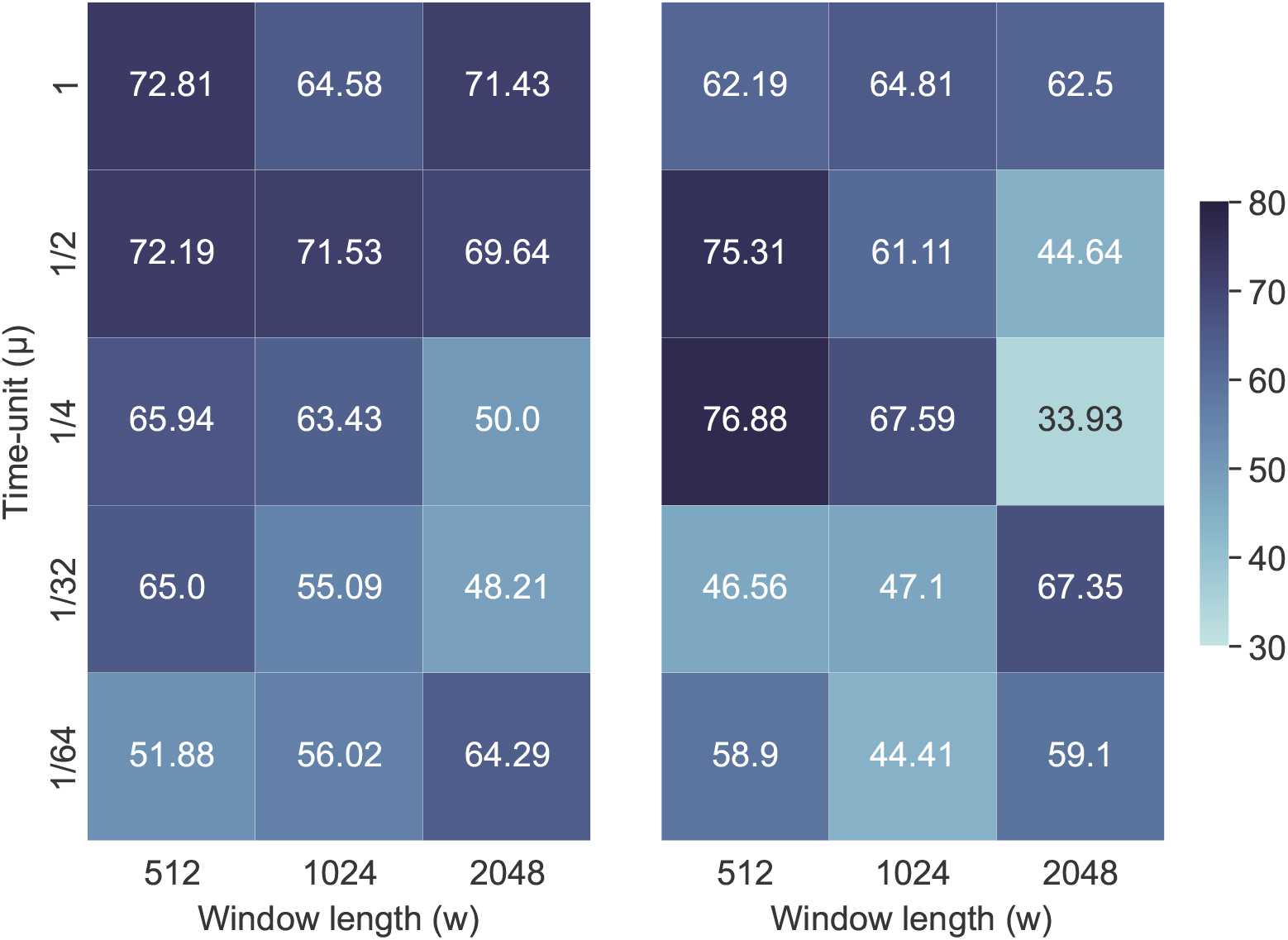
The validation accuracy of (Left) MLP and (Right) BiLSTM models trained on data with varying window size *w* and time-unit *µ*.

## 5 Discussion

As mental health has been lagging behind other areas of healthcare in the adoption of biomarkers to support clinical decision making, despite an urgent need thereof, the fortunate conjuncture of advances in machine learning and the wider availability of wearable devices and their improved precision to continuously and ecologically collect physiological data from patients holds great potential for the field. We herewith presented a pilot study in mood disorders and made the case that mood episode severity classification is a harder but clinically more relevant task in affective disorders (and mental health at large) than mere disease presence detection. We showcased a viable baseline for pre-processing raw data from the E4 Empatica, a popular research-grade wristband, for use in a machine learning pipeline and compared several algorithms, comprising both classical and deep-learning models, on mania and depression severity. Furthermore, when assessing model generalisation performance we set up two groups, one including segments from the same subjects used for model development and one of the segments from subjects the model had never seen during training. Overall, all models reached reasonable test performance in the former group, with DNNs leading by a comfortable margin over non-deep-learning models. However, when tested on the latter group, performance plunged for all models, the best one (BiLSTM) barely hovering above the random guessing threshold. Of note, DNNs were not any better here, with ENET (a very simple linear model) coming up on top of MLP. The poor performance of group 2, i.e. out-of-sample segments, can be expected considering the high inter-subjects heterogeneity: group 1 and group 2 patients were not exactly matched with each other for non-mood-disorders related (e.g. age, sex, level of physical fitness) or even some mood-disorders-related (e.g. administered treatment) variables. More to the point, this result also spells a well-described phenomenon in other applications of machine learning to mental health, e.g. neuro-imaging [46]. Non-disease-related dimensions of inter-patient variability, e.g. age, sex, physical health/fitness, tend to drown the disease-relevant signal so that, in our case, it can be fairly said that the model accurately picked up on individual-level patterns, as indicated by the high accuracy on hold-out samples from the same patients used for model development, but failed to learn affective-state-related patterns, generalisable across different individuals. Further to an obvious increase in the sample size for model development, so that the train set encompasses more heterogeneity and is more representative of the population we aim to model, this calls for *ad-hoc* techniques to disentangle inter-patient clinically irrelevant heterogeneity from the state-related signal.

### 5.1 Limitations

We acknowledge several limitations in our work. Most importantly, while the purpose of this work, as indeed a pilot, was exploratory, the small sample size available for model development did not allow us to make any strong claim about a machine learning system performance on new patients with manic or depressive episodes, unseen during model development. However, the recordings were all longer than 40 hours each, with fine-grained time-series collection of physiological variables ranging from 1 to 64Hz. Secondly, while converging evidence from other fields points to deep learning with raw (or minimally pre-processed) data as state of the art in time-series analysis, we will differ to future work on the investigation of performance with pre-defined features, extracted with human-designed tool-kits across all channels (in this study we only extracted features for IBI, since it does not lend itself to be time-aligned in its original format). Lastly, we appreciate that, while dealing with minimally pre-processed data, our pipeline can be improved in terms of artefacts detection and denoising (e.g. Kleckner et al. [27] and Taylor et al. [42]).

### 5.2 Future work

The pilot we herewith presented is the starting point of a broader project were, in general terms, we aim to harness machine learning and data from wearable devices to aid clinical decision making, empower patients, and advance the understanding of BD. We aim to collect data from over 60 patients with BD over the course of the first year.

Since psychiatry nosography almost entirely relies on clinical phenomenology and is not backed up by objective measures, the whole concept of ground truth in mental health diagnosis is brittle, with previous works indeed pointing to fairly low rates of inter-specialists agreement [45]. Thus, further to supervised learning in the context of disease detection and severity differentiation, a future line of research will be an unsupervised learning attempt at identifying disease groups based on data from E4 Empatica and validating such groups with respect to a clinically meaningful outcome. Secondly, as we are collecting detailed data on administered treatment, another future direction will be to find biomarkers from E4 Empatica data to predict treatment response or liability to side effects. All in all, to improve diagnosis and treatment in mental health from a precision psychiatry perspective.

## Data Availability

The anonymized raw recordings and psychometric scales used in this work will be made publicly available upon publication and completion of the funded project.

## Acknowledgement

We would like to thank Antonio Vergari for his insightful comments and suggestions on improving this manuscript. This research was supported by a grant from Baszucki Brain Research Fund and the Instituto de Salud Carlos III (FIS PI21/00340, TIMEBASE STUDY). Bryan M. Li and Filippo Corponi are supported by the United Kingdom Research and Innovation (grant EP/S02431X/1), UKRI Centre for Doctoral Training in Biomedical AI at the University of Edinburgh, School of Informatics. Gerard Anmella is supported by a Rio Hortega 2021 grant (CM21/00017) from the Spanish Ministry of Health financed by the Instituto de Salud Carlos III (ISCIII) and co-financed by the Fondo Social Europeo Plus (FSE+). Anna Giménez-Palomo thanks the support of an educational grant from the Spanish Ministry of Health, Instituto de Salud Carlos III (CM21/00094), and of the European Social Fund Plus (FSE+). Iria Grande thanks the support of the Spanish Ministry of Science and Innovation (MCIN) (PI19/00954) integrated into the Plan Nacional de I+D+I and cofinanced by the ISCIII-Subdirección General de Evaluación y el Fondos Europeos de la Unión Europea (FEDER, FSE, Next Generation EU/Plan de Recuperación Transformación y Resiliencia_PRTR); the Instituto de Salud Carlos III; the CIBER of Mental Health (CIBERSAM); and the the Secretaria d’Universitats i Recerca del Departament d’Economia i Coneixement (2017 SGR 1365), CERCA Programme / Generalitat de Catalunya. Diego Hidalgo-Mazzei’s research is supported by a Juan Rodés grant (JR18/00021) ISCIII.

## A Appendix

**Figure A.1:**
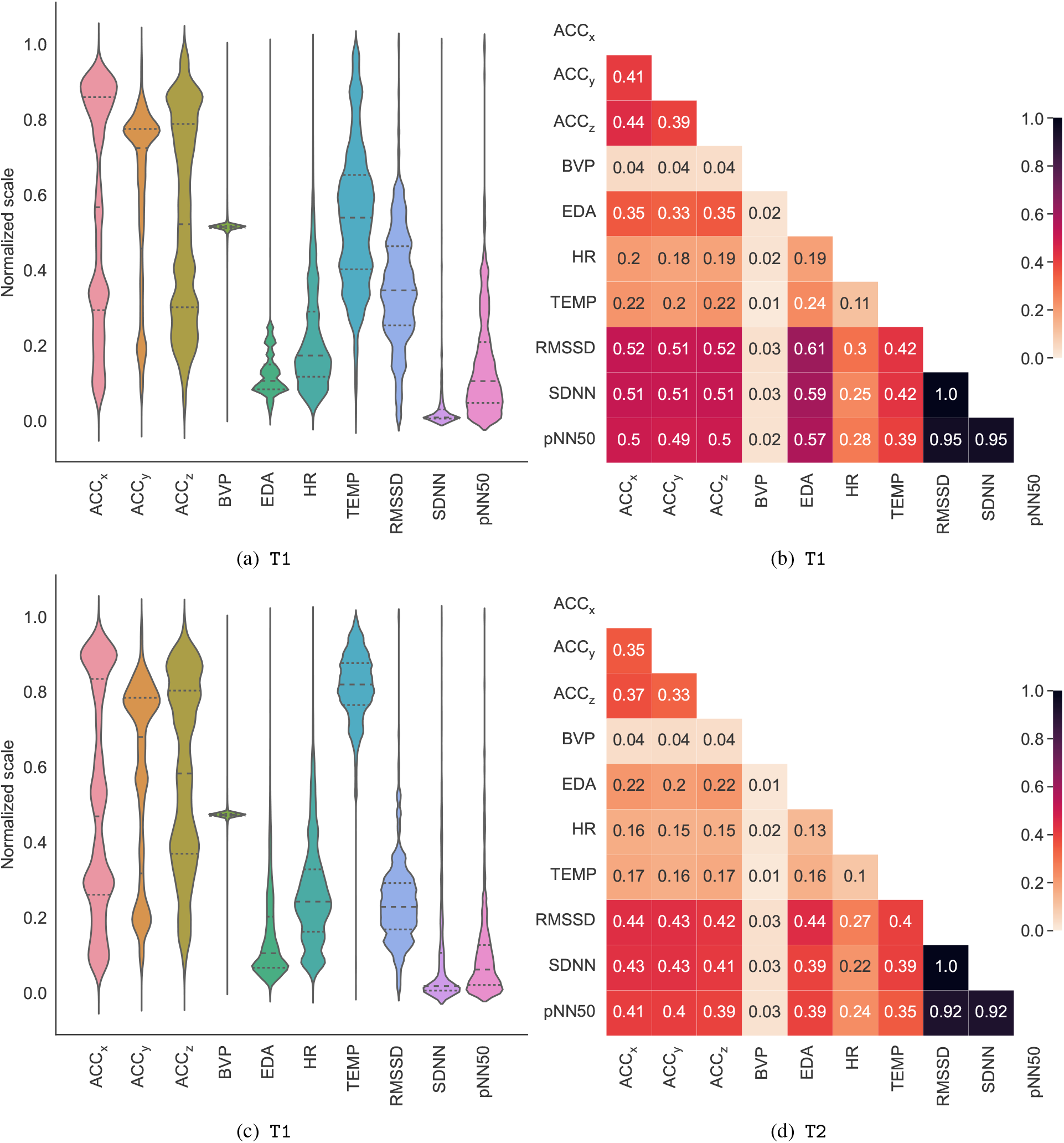
The (first column) per-channel normalised feature distributions and (second column) channel pairwise normalised mutual information of time-aligned recordings obtained from the subject on a major depression episode from group 1 at (first row) `T1` and (second row) `T2`.

**Table A.1:** The best model hyper-parameters after a random search on the dataset with window length *w* = 512 and time-unit *µ* = 1/2. α and l1_ratio_ denote the penalty term and mixing parameter in ENet. *C* is the regularisation parameter in SVC. The Gini impurity is being used as the criterion function in RF.

| Model | Configuration |
| --- | --- |
| ENET | $\alpha = 0.40049$ $l1_{\text{ratio}} = 0.00935$ |
| SVC | $C = 0.00409$ $\text{kernel} = \text{polynomial}$ |
| RF | $\text{num. estimators} = 560$ $\text{criterion} = \text{Gini}$ |
| MLP | $\text{num. layers} = 3$ $\text{layer size} = 128$<br>$\text{activation} = \text{LReLU}$ $\text{dropout} = 0.25$<br>$\text{optimiser} = \text{Adam}$ $\text{learning rate} = 0.001$ |
| BiLSTM | $\text{num. layers} = 1$ $\text{layer size} = 128$<br>$\text{activation} = \text{Tanh}$ $\text{recurrent activation} = \text{sigmoid}$<br>$\text{dropout} = 0$ $\text{recurrent dropout} = 0$<br>$\text{optimiser} = \text{Adam}$ $\text{learning rate} = 0.001$ |

**Figure A.2:**
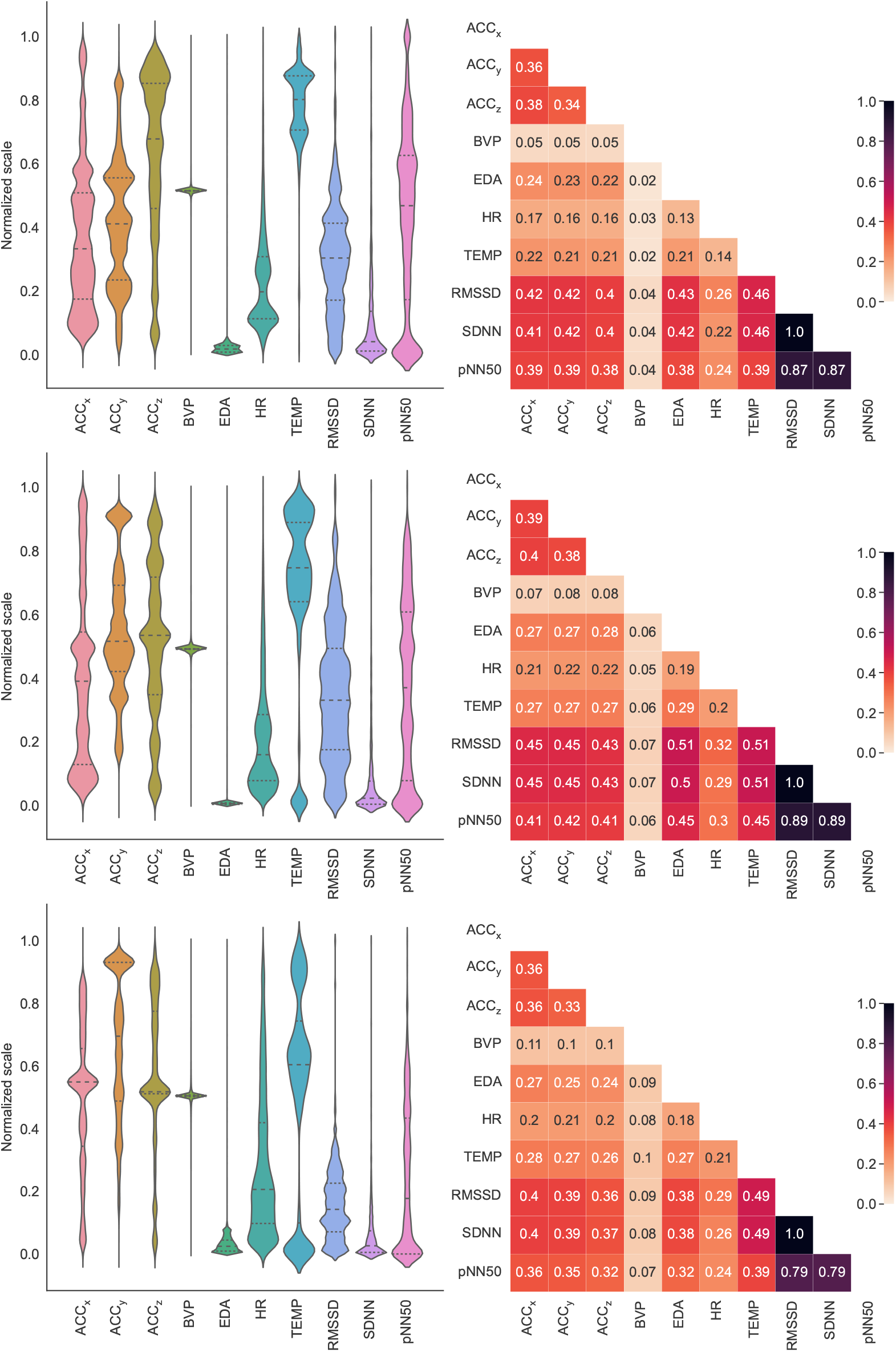
The (first column) per-channel normalised feature distributions and (second column) channel pairwise normalised mutual information of time-aligned recordings obtained from the subject on a manic episode from group 1 at (first row) `T0`, (second row) `T1` and (third row) `T2`.

**Figure A.3:**
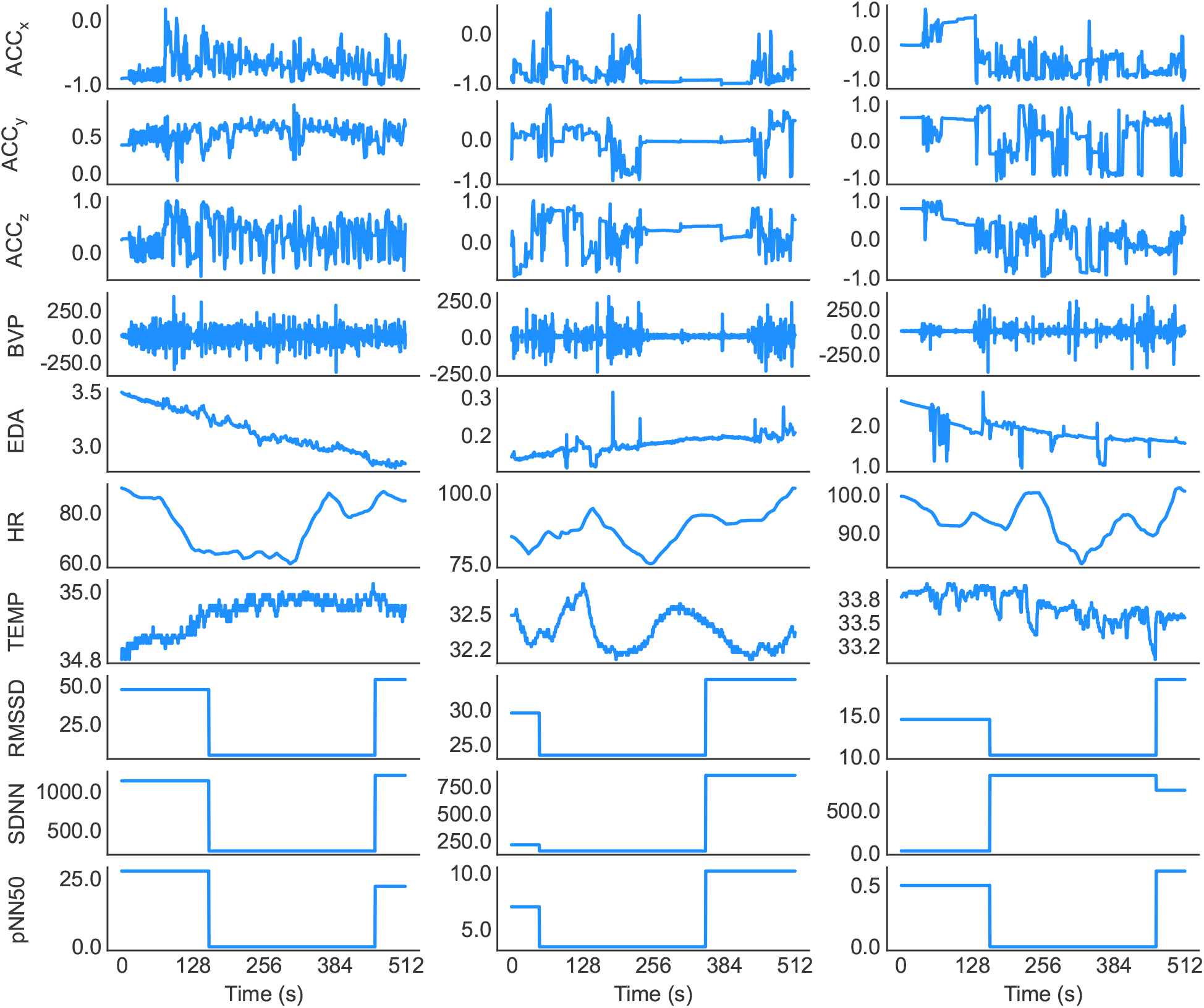
Randomly selected pre-processed segments of subject 1 (depressive) from Group 1 in (Left) `T0`, (Middle) `T1` and (Right) `T2` states (window length *w* = 512 and time-unit *µ* = 1/2). Note that the segments shown here have not been normalised and are in their original scale.

**Figure A.4:**
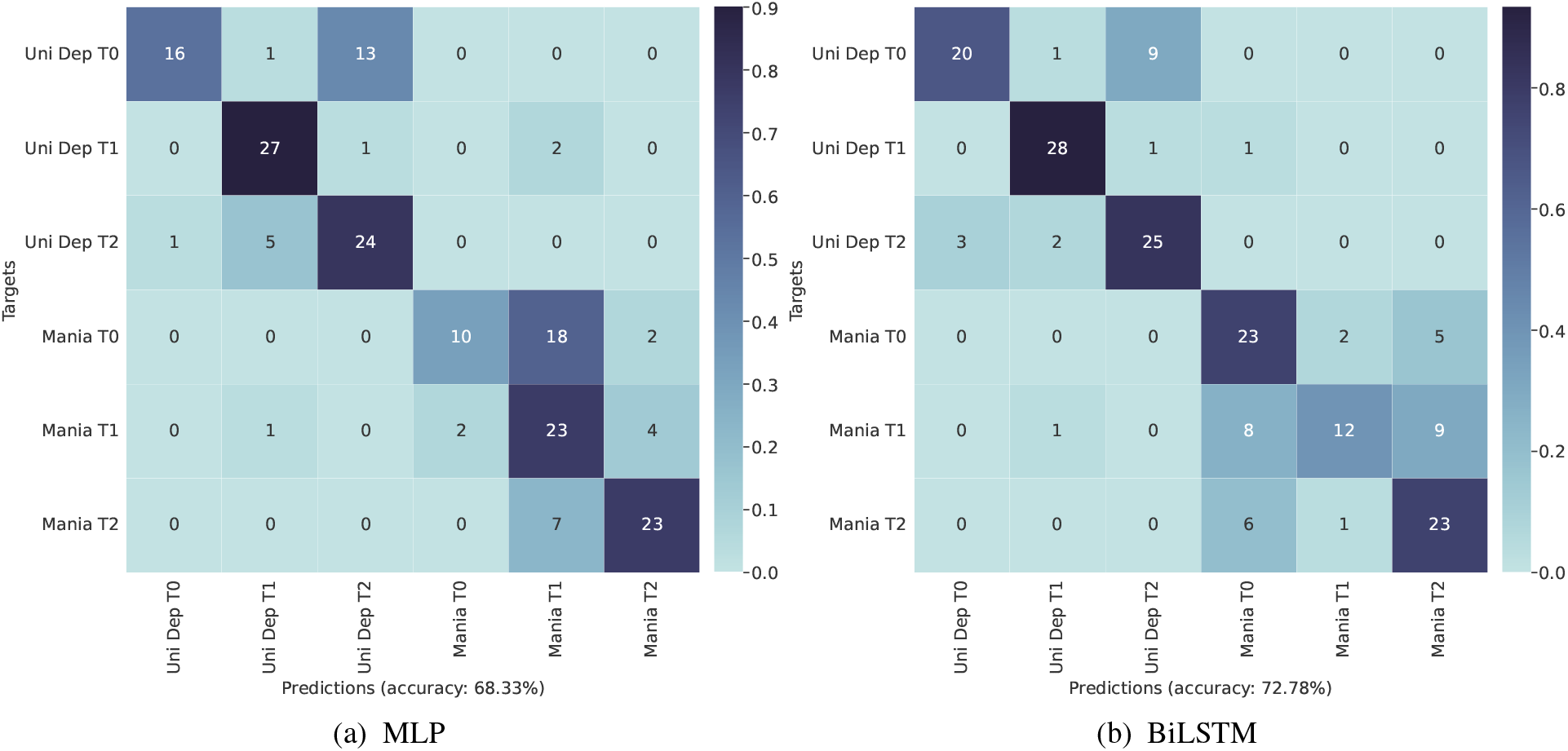
The classification confusion matrix of (a) MLP and (b) BiLSTM models on the group 1 test set (window length *w* = 512 and time-unit *µ* = 1/2).

## Footnotes

1 www.empatica.com/en-int/research/e4

2 https://github.com/Aura-healthcare/hrv-analysis

3 The software codebase will be made publicly available upon publication.

